# Comparison of Pericapsular Nerve Group and Lateral Quadratus Lumborum Blocks for Analgesia after Primary Total Hip Arthroplasty: A Randomized Controlled Trial

**DOI:** 10.1101/2024.07.18.24310628

**Authors:** Ellen L. H. Johnson, Tara L. Kelly, Bethany J. Wolf, Erik Hansen, Andrew Brown, Carla Lautenschlager, Sylvia H. Wilson

## Abstract

**Introduction:** The quadratus lumborum block (QLB) and the pericapsular nerve group (PENG) block both provide effective postoperative analgesia after hip surgery while minimizing impact on motor function. This study aimed to compare QLB and PENG in patients undergoing primary total hip arthroplasty.

**Methods:** This superiority trial randomized patients scheduled for elective total hip arthroplasty to receive a lateral QLB or PENG with lateral femoral cutaneous nerve blocks for postoperative analgesia. Perioperative analgesic protocols were standardized. The primary outcome was postoperative cumulative opioid consumption at 72 hours. Secondary outcome was postoperative pain scores. Additional outcomes of interest included time to first ambulation, length of stay, patient reported outcome measures, and opioid-related side effects.

**Results:** This trial consented and randomized 106 subjects and 101 were included in analysis: PENG (n=50), QLB (n=51). Mean (95% CI) opioid consumption (IV MME) in the first 72 hours did not differ between PENG [109.6 (93.6, 125.6)] and QL [92.3 (76.6, 107.9)] groups (p=0.129) There were no significant differences between treatment arms in average pain score, time to ambulation, distance ambulated, rate of same day discharge, or hospital length of stay. There were also no differences in patient reported outcomes using HOOS-JR and PROMIS-10 scores.

**Conclusion:** Patients undergoing primary THA receiving preoperative PENG vs QLB had similar opioid consumption, pain scores, time to ambulation, and hospital length of stay. Both QL and PENG blocks are analgesic options in patients undergoing primary THA.

**Clinical Trials Registration:** NCT05710107; www.ClinicalTrial.gov

IRB Protocol ID: Pro00124880

**Key message:**

- Pericapsular nerve group (PENG) block may provide analgesia after hip arthroplasty and improve early functional recovery. This study evaluated postoperative opioid consumption in patients randomized to PENG or lateral quadratus lumborum block (QLB).
- Opioid consumption, pain scores, motor recovery, and functional outcome measures did not differ in patients randomized to PENG vs lateral QLB.
- PENG and lateral QLBs are analgesic options following total hip arthroplasty with similar rates of same day discharge.

## INTRODUCTION

Total hip arthroplasty is a common orthopedic procedure and demand is expected to increase by 176% by 2040 and 659% by 2060(1). With this growth, interest in decreasing hospital length of stay has resulted in the development of rapid recovery protocols, including analgesia with regional anesthetic techniques promoting early ambulation(2, 3). The lateral quadratus lumborum block (QLB) has shown to be effective in controlling pain, increasing time to first request of analgesia, and decreasing opioid consumption after THA(4, 5), but few studies have compared the lateral QLB with other motor sparing blocks including the more recently described pericapsular nerve group (PENG) block.

The PENG block targets the articular branches of the obturator, accessory obturator, and femoral nerves, thus inhibiting sensory innervation to the anterior capsule while retaining motor innervation(6, 7). It can be combined with a lateral femoral cutaneous (LFC) nerve block to improve its analgesic effect on the lateral thigh. When compared to no block, PENG blocks have been shown to reduce pain scores, opioid use, and time to first ambulation while increasing hip range of motion(8). PENG blocks may also provide analgesia similar to fascia iliaca compartment blocks (FICB)(9) and improved analgesia compared to femoral nerve blocks(9), while maintaining quadriceps strength. However, the analgesic properties of PENG and lateral QLB have not been compared.

The purpose of this study was to compare postoperative opioid consumption in patients undergoing primary THA and randomized PENG and LFC blocks or a lateral QLB. We hypothesized that the combined PENG + LFC nerve block would provide superior analgesia, as measured by opioid consumption at 72 hours after surgery and pain scores, while promoting functional recovery.

## METHODS

This randomized trial was approved by the institutional review board (Pro00124880; 04/21/2020) and registered on www.ClinicalTrials.gov (NCT05710107; 1/12/2023) before patient enrollment. This trial was conducted in accordance with the original protocol, and written informed consent was obtained from all subjects. This manuscript adheres to the applicable Consolidated Standards of Reporting Trials guidelines.

On the day of surgery, subjects were invited to participate, provided with informed consent, and enrolled if eligible by study staff if determined to be eligible. Inclusion criteria included age ≥ 18 years of age and ambulatory patients undergoing elective hip arthroplasty with planned same day discharge or observation of 23 hours or less. Exclusion criteria included local anesthetic allergy, weight <40 kg, unable or unwilling to provide informed consent, and substance abuse. Enrollment and initial data collection took place at the Medical University of South Carolina University Hospital in Charleston, SC. Subsequent data was collected via a paper diary or secure text messaging system and in the orthopedic clinic.

Enrollment occurred from February 7, 2023 to November 9, 2023, with data collection continuing until December 16, 2023. Consenting subjects were consecutively assigned a study ID (1-106) and randomized to either lateral QLB, or PENG block with LFC block using a computer-generated list created by a statistician before study initiation using simple randomization. Randomization assignments were kept in sealed envelopes which were opened prior to block placement. Other than the regional anesthesia team, all patients, care team members, and research staff were blinded to randomization.

### Protocol

In preoperative holding, subjects were placed in the supine position and administered intravenous sedation (midazolam (0-2mg), dexmedetomidine (0-20μg)). To maintain blindness, all participants regardless of group assignment had sonographic scans of all block sites with aseptic skin prep and subcutaneous lidocaine placement at the appropriate insertion site. For both block groups, a 10 cm, 21-gauge, echogenic needle was inserted and ropivacaine (30mL, 0.25%) injected in 3-5 mL aliquots with intermittent aspiration.

### Lateral quadratus lumborum block (QL) Block

A high-frequency (13–6 MHz), linear or low-frequency (5–2 MHz), curvilinear ultrasound probe was used to visualize the external oblique, internal oblique, and transversus abdominus muscles before scanning laterally to identify the lateral aponeurosis of the transversus abdominus muscle and the lateral aspect of the QL muscle. An echogenic needle was then advanced (anterior to posterior) until the needle tip was deep to the aponeurosis to the transversalis abdominus muscle and lateral to the QL muscle, as previously described(10). Ropivacaine was injected incrementally with frequent aspiration and spread observed on ultrasound imaging.

### Pericapsular Nerve Group Block (PENG) Block with Lateral Femoral Cutaneous (LFC) Block

For PENG block placement, a low-frequency (5–2 MHz), curvilinear ultrasound probe was used to visualize the anterior inferior iliac spine, iliopsoas tendon, and iliopubic eminence. An echogenic needle was then advanced (lateral to medial) under ultrasound guidance until the tip reached the lateral and inferior margin of the iliopsoas tendon between the anterior inferior iliac spine (lateral) and iliopubic eminence (deep), as previously described. Ropivacaine (20ml; 0.25%) was injected lateral and inferior to the psoas tendon with spread observed on ultrasound along the lateral superior pubic ramus(7).

A high-frequency (13–6 MHz), linear probe was used to visualize the lateral femoral cutaneous (LFC) nerve superficial to the sartorius muscles and medial to the anterior superior iliac spine. An echogenic needle was then advanced (lateral to medial) under ultrasound guidance until the needle tip was visualized in plane with the nerve(11). Ropivacaine (10ml; 0.25%) was injected and spread observed around the nerve.

### Anesthetic Care

Anesthetic care was standardized. Unless contraindicated, multimodal analgesia included preoperative oral acetaminophen (1000mg) and intraoperative ketorolac (15–30 mg intravenous, IV, based on renal function) during surgical closure. Intraoperatively, spinal anesthesia (bupivacaine 10mg) was supplemented with intravenous sedation. General anesthesia was utilized in the event of the inability to place or inadequate spinal anesthesia. Surgeons performed periarticular injection immediately before closure (ropivacaine 0.2% with ketorolac 30 mg and clonidine 100mcg) for all patients. Postoperative anesthetic care unit (PACU) orders were standardized and included hydromorphone (0.2mg IV every 10min for severe pain). Postoperative surgical orders were standardized: oral acetaminophen (1000mg every 8hours), methocarbamol (750–1000mg every 8hours), celecoxib (200mg two times per day), and oxycodone as needed for moderate and severe pain (5–10 mg).

### Outcomes

Collected data included demographics, opioid consumption, pain rating using the Visual Analog Scale (VAS), time to first ambulation (time 0 with operative spinal placement), distance at first ambulation, post anesthesia care unit (PACU) duration, hospital length of stay (LOS), same day discharge rates, patient reported outcome measures [Hip disability and Osteoarthritis Outcome Score for Joint Replacement (HOOS JR) and Patient-Reported Outcome Measures Information System (PROMIS-10) surveys], and opioid related side effects. Demographic data collected included patient age, sex, and race, and body mass index (BMI, cm^2^/kg). Intraoperative and postoperative opioids were converted to intravenous morphine mg equivalents (IV MME) for comparison. VAS measurements were taken by having patients mark on a 100mm line (0mm: no pain to 100mm: worst pain). VAS measurements were taken before placing the block in holding and approximately one hour after PACU arrival. Starting on postoperative day (POD) 1, data was collected by paper pain diary or by secure text message (Twilio(12)) based on the patient’s preference. Specifically, the name, dose, and frequency of any pain medications, VAS measurements, patient satisfaction, and any side effects were collected. For subjects preferring paper diaries, a member of the research staff would call and speak to patients for data collection. For subjects preferring texts, paper diaries were also given, and messages were sent at 9 am (to collect overnight data) and 3 pm (to collect day data) through POD 3. PACU duration, hospital length of stay, and same day discharge rates were collected by research staff. Time to first ambulation and distance ambulated were collected by physical exam by trained research staff in PACU and by physical therapy assessments. On POD 7, 2 weeks postoperative, and 6 weeks postoperative, participants completed the HOOS Jr and PROMIS-10 Global Health questionnaires by text message or phone interview from the research team. HOOS JR provides information on patient-reported hip pain and function in patients undergoing THA(13). PROMIS-10 provides information on patient-reported general health quality of life, including physical, mental, and social health. Opioid related side effects were collected throughout the study.

### Power

Postoperative cumulative opioid consumption (IV MME) at 72 hours was the primary outcome. A prior study(10) found mean IV MME at 12 hours postoperative in subjects undergoing THA with a QL block was 16 ±12 MME. Therefore, a sample size of 48 subjects per group with 9 repeated measures of opioid consumption provides 80% power to detect a difference in cumulative MMEs consumed postoperatively of 4 MMEs at significance level α = 0.006 (Bonferroni adjusted for 9 pairwise comparisons between group) assuming a standard deviation of ±12 MMEs, a first-order autoregressive correlation structure and correlation between observations on the same subject of ρ = 0.33. Thus, enrollment planned for 53 subjects/group (106 total) to allow for 10% attrition.

### Statistics

Differences in patient characteristics between treatment arms were assessed using chi-square or Fisher’s exact test for categorical variables and t-tests or Wilcoxon rank sum tests for continuous variables to assess balance between treatment arms.

The primary outcome of interest was cumulative postoperative opioid consumption in the first 72 hours. Differences in cumulative opioid consumption between block groups was assessed using a linear mixed model approach. The model included fixed effects for treatment group, postoperative time, and the interaction between block group and postoperative time and a random subject effect to account for correlation between measures collected on the same patient over time. Model assumptions were checked graphically, and transformations were considered if needed. Differences between block types at each postoperative time were evaluated using linear contrasts from the model.

Secondary and additional outcomes of interest included postoperative worst and average VAS pain scores in the first 72 hours, time to first ambulation, PACU duration, hospital length of stay, same day discharge rates, patient reported outcome measures (HOOS JR and PROMIS-10), and opioid related side effects. VAS pain score, patient reported outcomes (HOOS JR and PROMIS-10) were measured at multiple timepoints. Differences in these outcomes over time between block groups were assessed using a linear mixed model approach described previously. Difference between treatment groups in time to first ambulation, time to return of motor function, PACU duration, and hospital length of stay were evaluated using Wilcoxon rank sum tests. Associations between block type and opioid side effects were evaluated using Fisher’s exact test. The association between block type and rate of same day discharge was evaluated using a logistic regression approach. The model included block type and anesthesia stop time to account for the fact that later surgeries were less likely to go home the same day.

Approximately 0-21% of observations for primary and secondary outcomes were missing. Multiple imputation with 10 imputations was used to impute missing outcome values prior to all analyses and results are reported based on the pooled estimates across imputations. As a sensitivity analysis, we also conducted a complete case analysis and compared the results with those from the imputed data. All analyses were conducted in SAS v. 9.4 (SAS Institute, Cary, NC, USA).

## RESULTS

This trial consented and randomized 106 subjects (February 7, 2023-November 12, 2023). Five patients were excluded from analysis due to withdrawal of participation (n=3, 2 in QLB group, 1 in PENG group) and inability to perform block (n=2, both in PENG group). The final study population included 50 subject who received a PENG block and 51 subjects who received a QL block (Figure 1). Participant characteristics by treatment group are shown in Table 1. There were no notable differences between demographic and baseline characteristics of the two groups.

**Figure 1.**
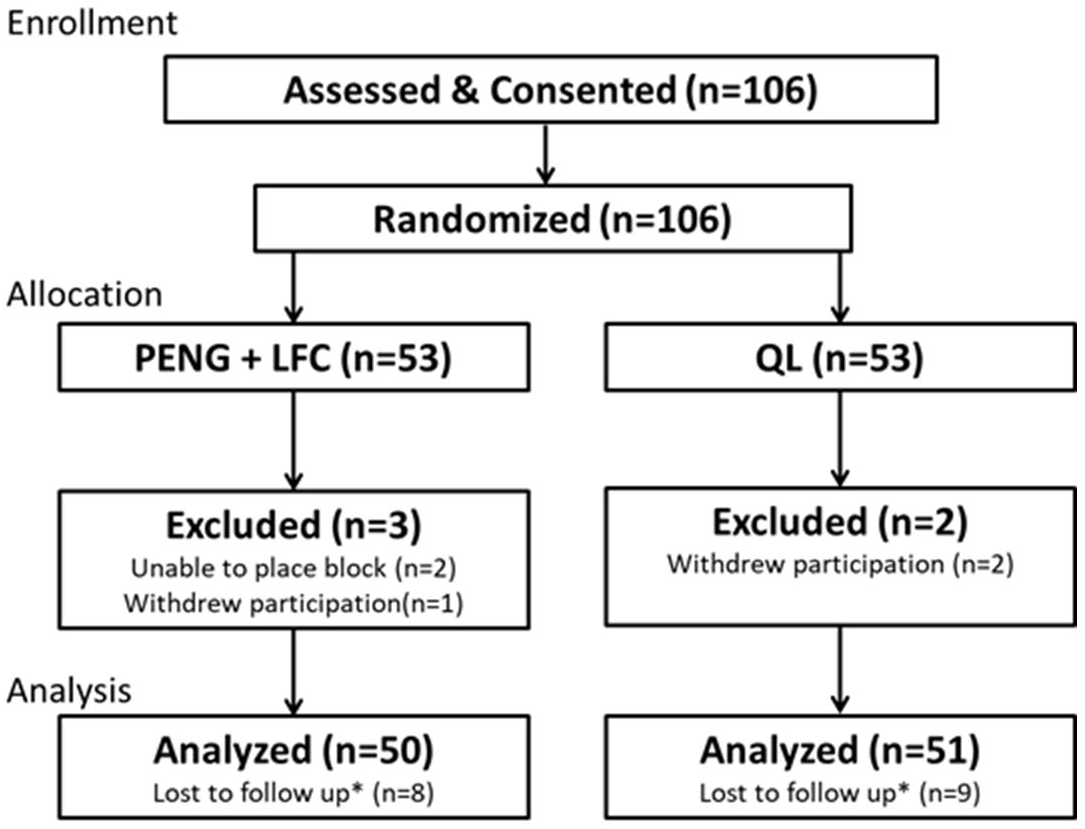
Consolidated Standards of Reporting Trials flow diagram. LFC, lateral femoral cutaneous nerve; PENG, pericapsular nerve group; QLB, quadratus lumborum block.

**Table 1.**
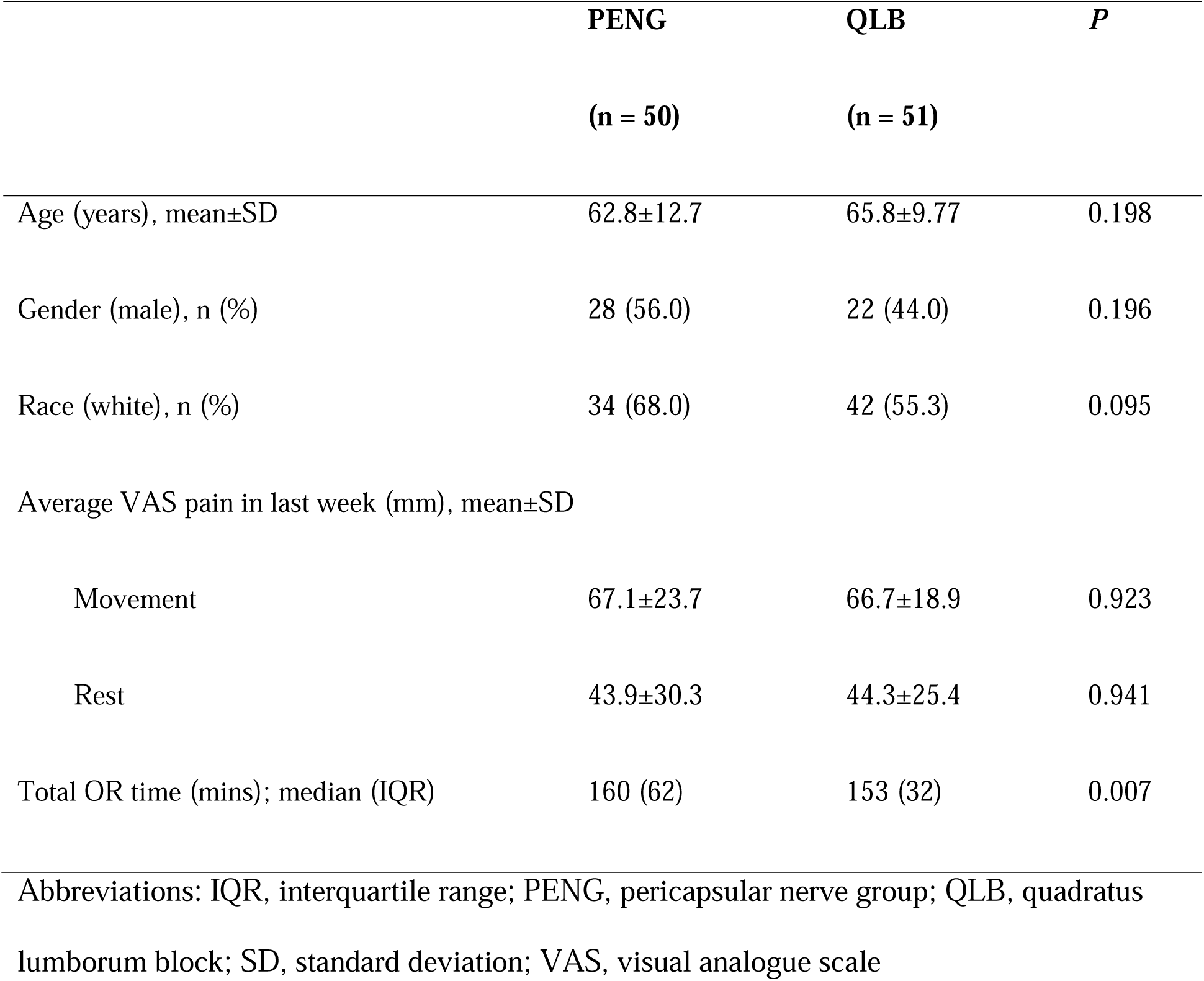
Patient Characteristics.

### Postoperative Opioid Consumption

Cumulative opioid consumption is presented in Figure 2. Mean (95% CI) opioid consumption (IV MME) in the first 72 hours did not differ between patients randomized to PENG [112.9 (93.4, 132.4)] and QLB [89.3 (71.1, 107.9); p=0.065]. A complete case analysis was conducted to serve as a sensitivity analysis to examine the impact of imputation to address missing data and results were similar.

**Figure 2.**
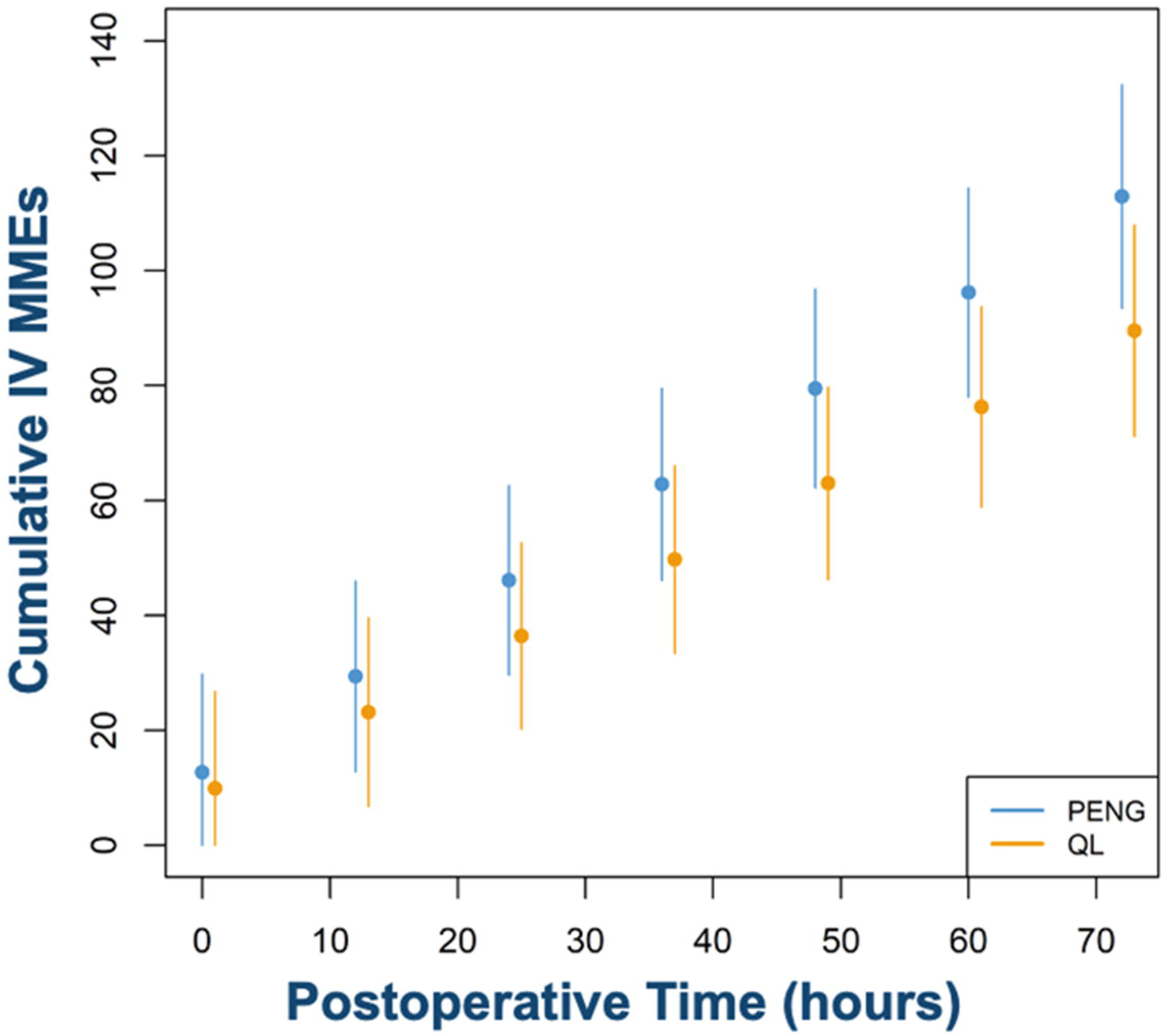
Cumulative Opioid Consumption.

### Additional Outcomes

Additional outcomes of interest are summarized in Table 2. There was no significant difference in average VAS pain scores between treatment arms. Worst pain reported on a VAS scale was an average of 7mm higher in the PENG group compared to the QLB group (p=0.032); however, when using imputed data, this difference was no longer significant when considering the complete case (p=0.061). There was no difference between treatment arms in time to first ambulation or distance ambulated. There was also no difference in patient centered outcome measures using HOOS and PROMIS scores at 1, 2, and 6 weeks postoperative.

**Table 2.**
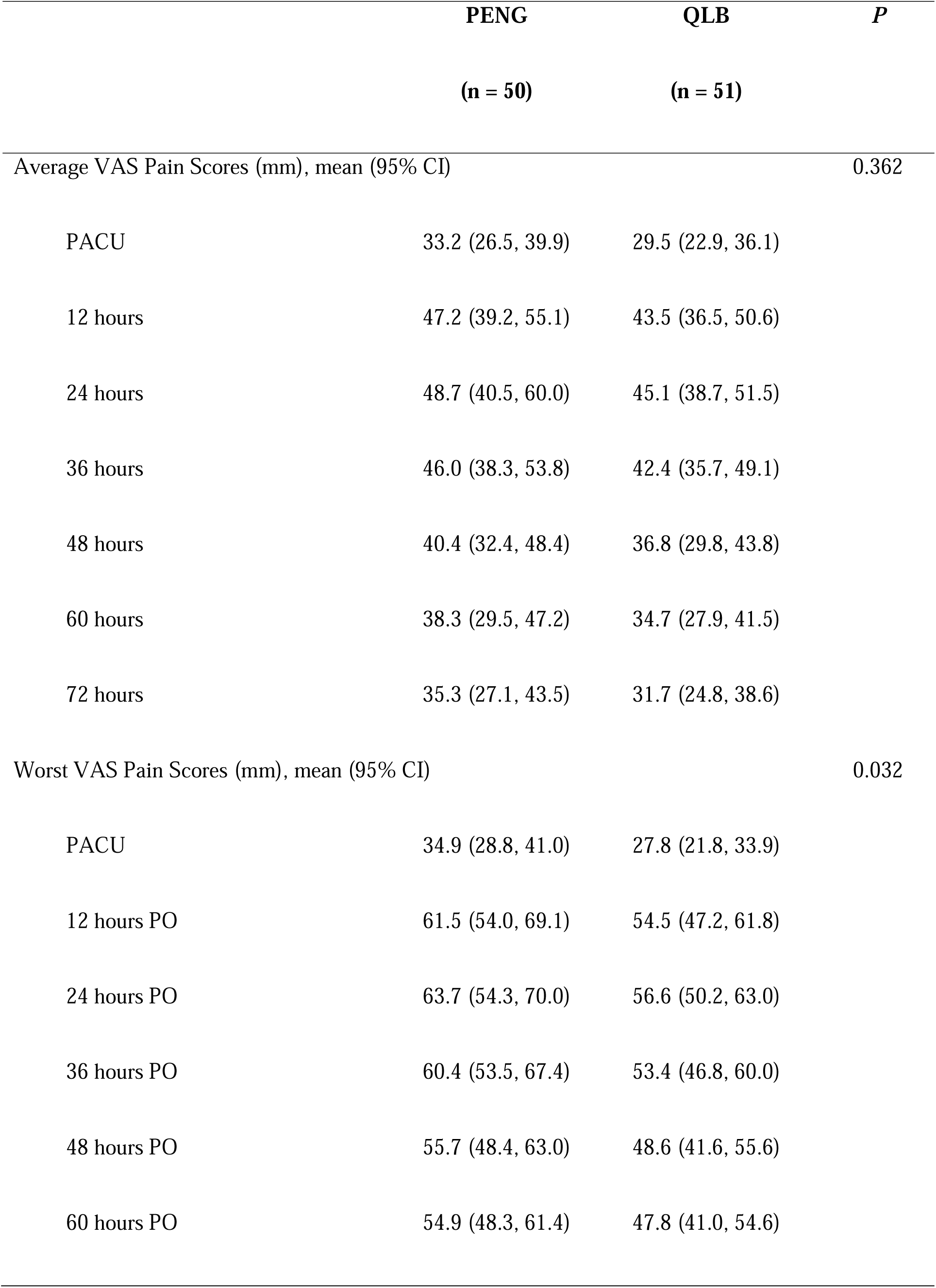
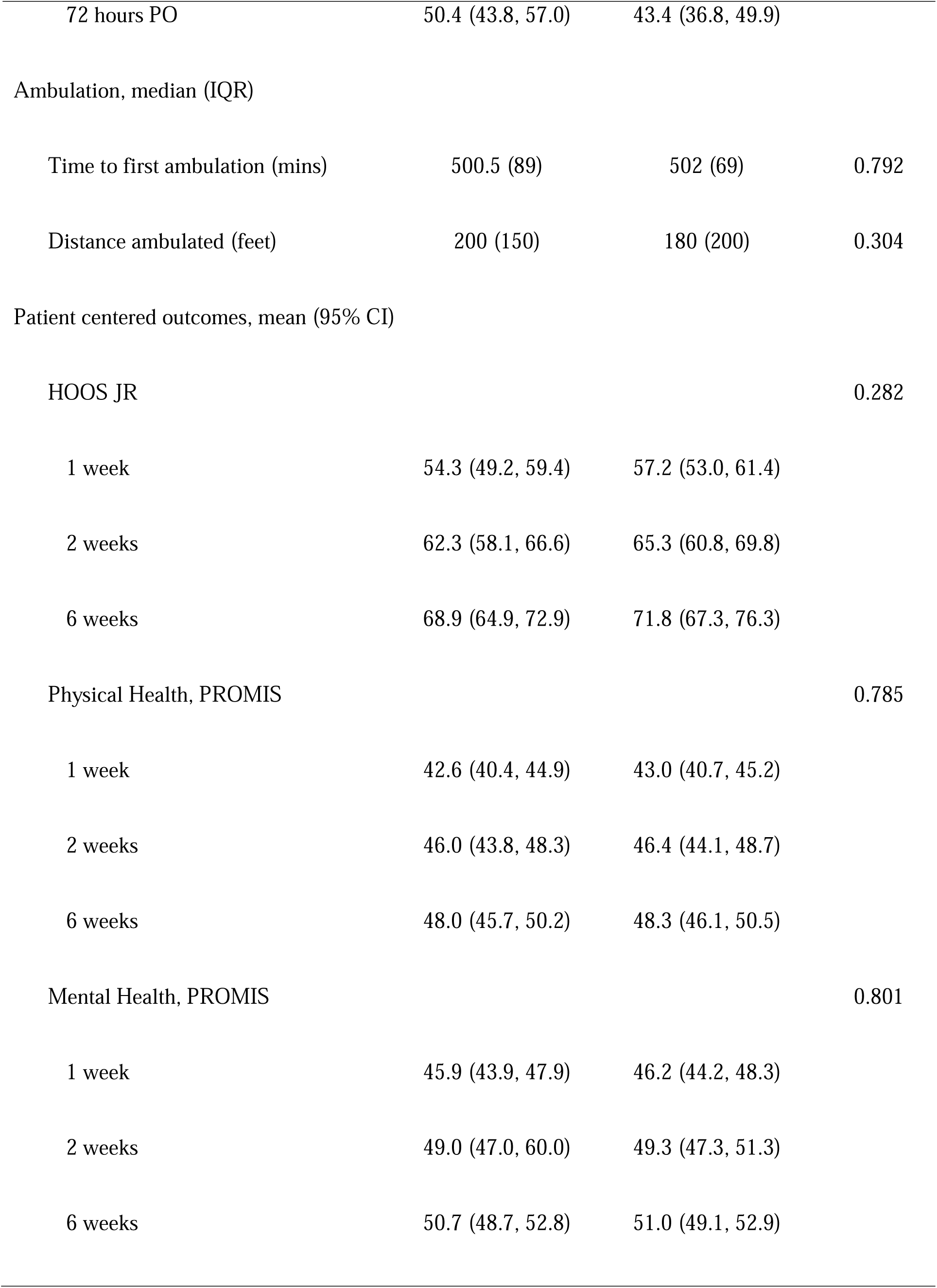
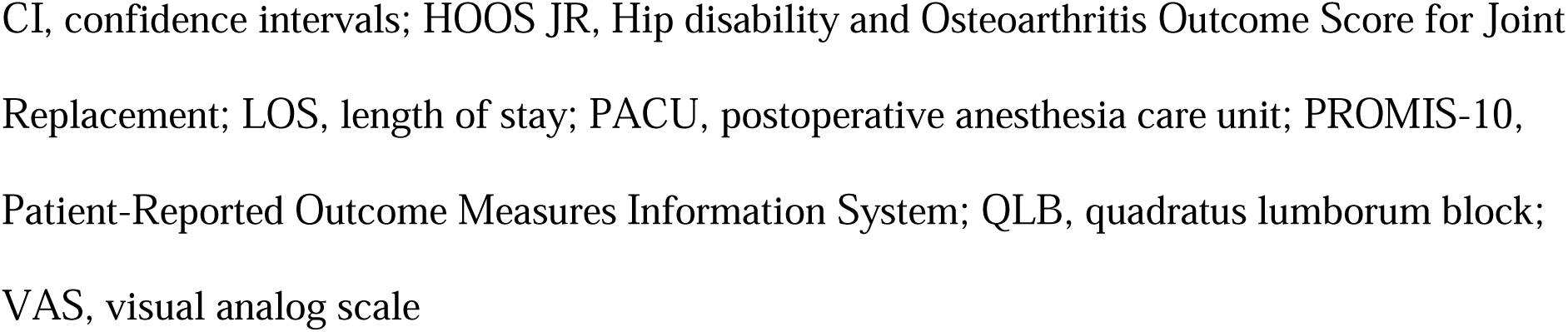
Additional Outcomes of Interest.

Hospital LOS is presented in Figure 3. Mean differences (95% CI) in PENG versus QLB groups for PACU LOS [-19 min (-74.0, 37.0); *P*=0.486) and hospital LOS [0.15 hours (-1.3, 2.05), *P*=0.821] and rates of same day discharge (48% versus 52%, respectively; *P*=0.561) did not differ between groups. Opioid side effects did not differ between groups in the first 72 hours including any (*P*=0.366), itching (*P*=0.352), and nausea (*P*=0.145). There was no documented harm or unintended effect in either group.

**Figure 3.**
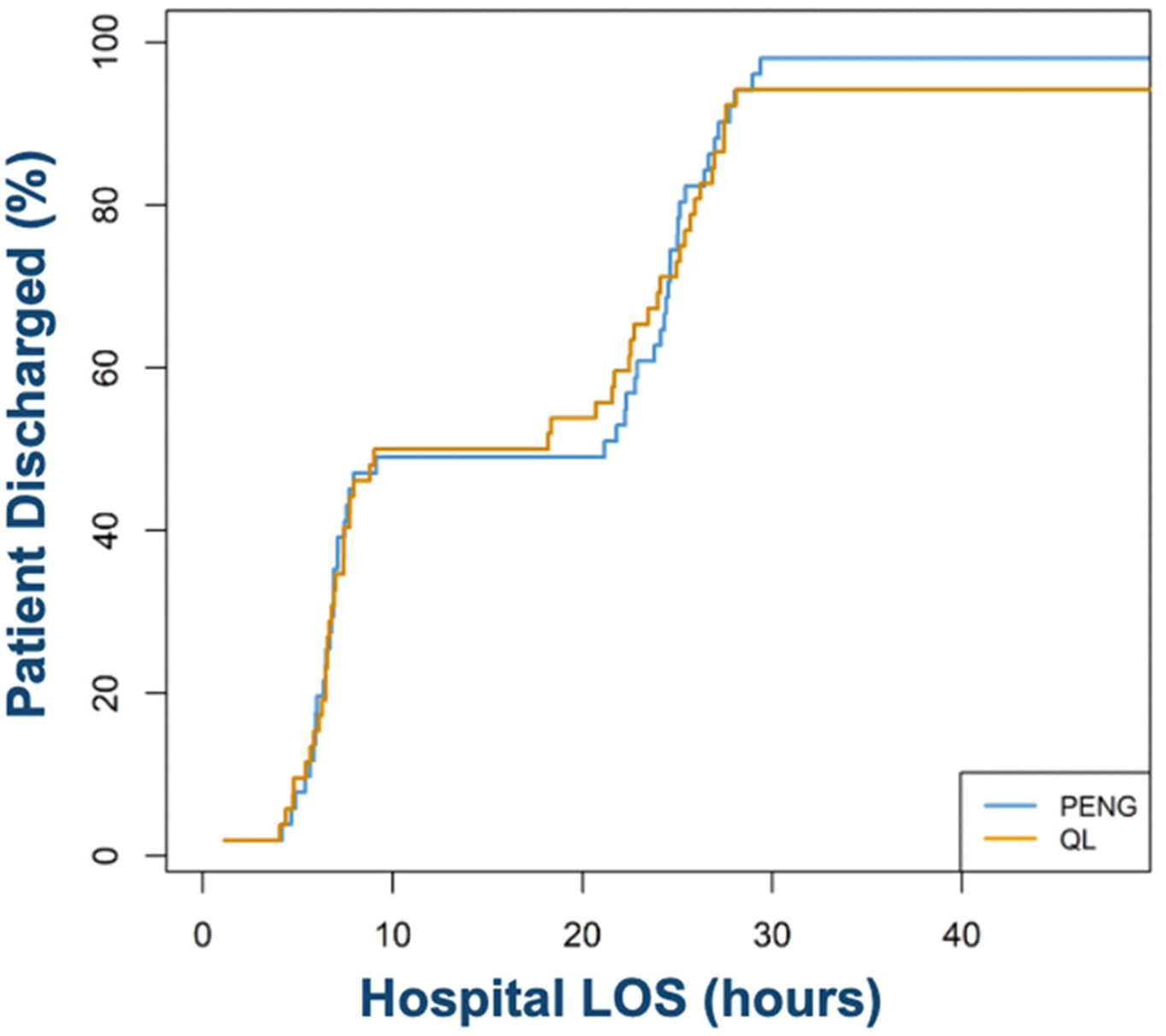
Patients Discharged Over Time.

## DISCUSSION

This randomized, prospective study did not find a PENG block with LFC block to be superior to a lateral QLB for reducing postoperative opioid consumption or pain scores after THA. Our results contrast with two prior studies which found patients randomized to PENG for THA (14) or surgery for hip fracture (15) to have decreased opioid consumption compared to those receiving a fascia iliaca compartment block (FICB). Alternatively, our results support prior retrospective and smaller studies finding no difference in opioid consumption after PENG compared to other peripheral nerve blocks. (9, 16–19) In a retrospective study of 160 patient receiving a PENG (n=45), anterior QLB (n=38), or no block (n=77) for analgesia after THA, both peripheral nerve blocks were associated with decreased opioid consumption compared to controls, but opioid consumption did not differ between patient who received PENG or QLB (16). Similarly, in a prospective study of 89 patients for THA randomized to PENG (n=30), anterior QLB (n=30), or intraarticular injection (n=29), opioid consumption at 48 hours differed between the QLB and intraarticular but not between the PENG and other groups (18). Opioid consumption after THA also did not differ in a third trial of 90 patients randomized to PENG or anterior QLB(19) or in two recent prospective studies of 40 (9) and 58 (17) patients randomized to PENG or FICB. Our study also did not find a difference in pain score between block groups. While prior publications noted no differences in pain scores when comparing PENG to anterior QLB (18) or FICB (20), other studies have noted improved pain scores with PENG compared with anterior QLB (19), FICB (14, 15, 17), and femoral neve block (21). While our findings support that both PENG and latera QLB offer analgesia following THA, they also support the use of the lateral QLB to promote early functional recovery including ambulation and same day discharge.

As joint arthroplasty increasingly moves to an outpatient procedure, earlier ambulation coupled with analgesia promotes same day discharge and peripheral nerve block selection may impact this goal. In a prospective study, patients randomized to PENG had similar quadriceps strength compared to patient with a sham block, but demonstrated earlier postoperative mobility with longer distances ambulated and earlier discharge home (22). Other publications have found patients randomized to PENG to have less quadriceps weakness compared with FICB (9), femoral nerve block (21), and anterior QLB (18). However, this improved strength and mobilization has not always correlated with a decreased LOS(8, 9, 19, 20). Our results support prior publications demonstrating that PENG blocks do not create quadriceps weakness and should promote the ability of patients to achieve same day discharge goals after THA but that LOS is likely multifactorial.

Fewer studies have evaluated functional outcomes measures after PENG. In 90 patients randomized the PENG or sham for THA, PENG subjects had improved quality of recovery (QoR)-15 scores on postoperative day 1 and 2 but this difference was lost on postoperative day 3 (22). Similar to our findings, QoR-40 scores did not differ in patients randomized to PENG, anterior QL, and intraarticular injection after THA, despite decreased pain and opioid consumption in both peripheral block groups(18).

Finally, our results are generalizable to most patients undergoing elective THA. We had few exclusion criteria and included most of our patient population presenting for primary THA.

### Limitations

Our study does have some potential limitations. While our group performs both PENG and QLB daily, PENG is a new technique. Notably, we were unable to perform the randomized block only in two patients, both randomized to the PENG group, which may suggest a more difficult block technique. Despite substantial institutional progress to increase patient ambulation after surgery, our physical therapists frequently do not start postoperative day zero ambulation until after noon, regardless of patient strength and readiness, in an effort to expedite discharge of any patients that stayed overnight for 23-hour observation. Thus, patient strength/readiness and pain control may not have been the primary factor impacting time or first ambulation. Similarly, while 88.15% of our total joint arthroplasty cases were outpatient in our institution in 2024, only 51.53% completed same day discharge with the patient/family preference being the primary reason for not going home on the day of surgery. Thus, patient/family choice and preparedness like remains a key factor in the ability to achieve same day discharge.

## Conclusion

Although we were unable to demonstrate superiority for opioid consumption at 72 hours postoperative, there was no significant difference in opioid consumption between groups. Additionally, mobility and functional outcome measures were similar between groups. We propose that both a lateral QLB and PENG block + LFC block are effective analgesic methods for patients undergoing THA.

## Data Availability

All data produced in the present work are contained in the manuscript

## Contributor statements

1. Ellen L. H. Johnson. Conflicts of interest: none. This author helped with study design, data collection, interpretation of the results, and manuscript writing and editing.
2. Tara L. Kelly. Conflicts of interest: none. This author helped with data collection, interpretation of the results, and manuscript writing and editing.
3. Bethany J. Wolf. Conflicts of interest: none. This author helped with study design, data interpretation, statistical analysis, and manuscript writing and editing.
4. Erik Hansen. Conflicts of interest: This author helped with study design, interpretation of the results, and manuscript writing and editing.
5. Andrew Brown. Conflicts of interest: none. This author helped with data collection and manuscript writing and editing.
6. Carla Lautenschlager. Conflicts of interest: none. This author helped with data collection and manuscript writing and editing.
7. Sylvia H. Wilson. Conflicts of interest: none. This author helped with study conception, study procedures, interpretation of the results, and manuscript writing and editing.

## Prior Presentation

This work was presented at The Society for Ambulatory Anesthesia 2024 Annual Meeting (Savannah, GA)

## Abbreviations

BMI: body mass index
CI: confidence intervals
EOM: external oblique muscle
FICB: facia iliaca compartment block
HOOS JR: Hip disability and Osteoarthritis Outcome Score for Joint Replacement
IOM: internal oblique muscle
IQR: interquartile range
IV MME: intravenous morphine mg equivalents
LDM: latissimus dorsi muscle
LOS: length of stay
PACU: postoperative anesthesia care unit
PROMIS-10: Patient-Reported Outcome Measures Information System
QL: quadratus lumborum
QLB: quadratus lumborum block
SD: standard deviation
THA: total hip arthroplasty
TAM: transversus abdominus muscle
VAS: visual analog scale.

## REFERENCES

1. Pabinger C, Lothaller H, Portner N, Geissler A. Projections of hip arthroplasty in OECD countries up to 2050. Hip Int. 2018;28(5):498–506.

2. Guerra ML, Singh PJ, Taylor NF. Early mobilization of patients who have had a hip or knee joint replacement reduces length of stay in hospital: a systematic review. Clin Rehabil. 2015;29(9):844–54.

3. Tasso F, Simili V, Di Matteo B, Monteleone G, Martorelli F, De Angelis A, et al. A rapid recovery protocol for hip and knee replacement surgery: a report of the outcomes in a referral center. Eur Rev Med Pharmacol Sci. 2022;26(10):3648–55.

4. Huda AU, Minhas R. Quadratus Lumborum Block Reduces Postoperative Pain Scores and Opioids Consumption in Total Hip Arthroplasty: A Meta-Analysis. Cureus. 2022;14(2):e22287.

5. Kim YJ, Kim HT, Kim HJ, Yoon PW, Park JI, Lee SH, et al. Ultrasound-Guided Anterior Quadratus Lumborum Block Reduces Postoperative Opioid Consumption and Related Side Effects in Patients Undergoing Total Hip Replacement Arthroplasty: A Propensity Score-Matched Cohort Study. J Clin Med. 2021;10(20).

6. Del Buono R, Padua E, Pascarella G, Costa F, Tognu A, Terranova G, et al. Pericapsular hip radiofrequency: future approaches to treat hip chronic pain. Minerva Anestesiol. 2021;87(12):1393–4.

7. Giron-Arango L, Peng PWH, Chin KJ, Brull R, Perlas A. Pericapsular Nerve Group (PENG) Block for Hip Fracture. Reg Anesth Pain Med. 2018;43(8):859–63.

8. Pascarella G, Costa F, Del Buono R, Pulitano R, Strumia A, Piliego C, et al. Impact of the pericapsular nerve group (PENG) block on postoperative analgesia and functional recovery following total hip arthroplasty: a randomised, observer-masked, controlled trial. Anaesthesia. 2021;76(11):1492–8.

9. Aliste J, Layera S, Bravo D, Jara A, Munoz G, Barrientos C, et al. Randomized comparison between pericapsular nerve group (PENG) block and suprainguinal fascia iliaca block for total hip arthroplasty. Reg Anesth Pain Med. 2021;46(10):874–8.

10. Kelly T, Wolla CD, Wolf BJ, Hay E, Babb S, Wilson SH. Comparison of lateral quadratus lumborum and lumbar plexus blocks for postoperative analgesia following total hip arthroplasty: a randomized clinical trial. Reg Anesth Pain Med. 2022;47(9):541–6.

11. Davies A, Crossley A, Harper M, O’Loughlin E. Lateral cutaneous femoral nerve blockade-limited skin incision coverage in hip arthroplasty. Anaesth Intensive Care. 2014;42(5):625–30.

12. Twilio. n.d. “What is Twilio, and how does it work? An introduction to the leader in customer engagement” https://www.twilio.com/en-us/resource-center/what-is-twilio-an-introduction-to-the-leading-customer-engagement-platform.

13. Jacobs CA, Peabody MR, Duncan ST, Muchow RD, Nunley RM, Group A, et al. Development of the HOOS(global) to Assess Patient-Reported Outcomes in Patients Undergoing Hip Preservation Procedures. Am J Sports Med. 2018;46(4):940–6.

14. Vamshi C, Sinha C, Kumar A, Kumar A, Kumari P, Kumar A, et al. Comparison of the efficacy of pericapsular nerve group block (PENG) block versus suprainguinal fascia iliaca block (SFIB) in total hip arthroplasty: A randomized control trial. Indian J Anaesth. 2023;67(4):364–9.

15. Mosaffa F, Taheri M, Manafi Rasi A, Samadpour H, Memary E, Mirkheshti A. Comparison of pericapsular nerve group (PENG) block with fascia iliaca compartment block (FICB) for pain control in hip fractures: A double-blind prospective randomized controlled clinical trial. Orthop Traumatol Surg Res. 2022;108(1):103135.

16. Braun AS, Peabody Lever JE, Kalagara H, Piennette PD, Arumugam S, Mabry S, et al. Comparison of Pericapsular Nerve Group (PENG) Block Versus Quadratus Lumborum (QL) Block for Analgesia After Primary Total Hip Arthroplasty Under Spinal Anesthesia: A Retrospective Study. Cureus. 2023;15(12):e50119.

17. Choi YS, Park KK, Lee B, Nam WS, Kim DH. Pericapsular Nerve Group (PENG) Block versus Supra-Inguinal Fascia Iliaca Compartment Block for Total Hip Arthroplasty: A Randomized Clinical Trial. J Pers Med. 2022;12(3).

18. Et T, Korkusuz M. Comparison of the pericapsular nerve group block with the intra-articular and quadratus lumborum blocks in primary total hip arthroplasty: a randomized controlled trial. Korean J Anesthesiol. 2023;76(6):575–85.

19. Wang QR, Ma T, Hu J, Yang J, Kang PD. Comparison between ultrasound-guided pericapsular nerve group block and anterio quadratus lumborum block for total hip arthroplasty: a double-blind, randomized controlled trial. Eur Rev Med Pharmacol Sci. 2023;27(16):7523–32.

20. Bravo D, Aliste J, Layera S, Fernandez D, Erpel H, Aguilera G, et al. Randomized clinical trial comparing pericapsular nerve group (PENG) block and periarticular local anesthetic infiltration for total hip arthroplasty. Reg Anesth Pain Med. 2023;48(10):489–94.

21. Lin DY, Morrison C, Brown B, Saies AA, Pawar R, Vermeulen M, et al. Pericapsular nerve group (PENG) block provides improved short-term analgesia compared with the femoral nerve block in hip fracture surgery: a single-center double-blinded randomized comparative trial. Reg Anesth Pain Med. 2021;46(5):398–403.

22. Hu J, Wang Q, Hu J, Kang P, Yang J. Efficacy of Ultrasound-Guided Pericapsular Nerve Group (PENG) Block Combined With Local Infiltration Analgesia on Postoperative Pain After Total Hip Arthroplasty: A Prospective, Double-Blind, Randomized Controlled Trial. J Arthroplasty. 2023;38(6):1096–103.

